# Longitudinal monitoring of SARS-CoV-2 RNA on high-touch surfaces in a community setting

**DOI:** 10.1101/2020.10.27.20220905

**Authors:** Abigail P. Harvey, Erica R. Fuhrmeister, Molly Cantrell, Ana K. Pitol, Jenna M. Swarthout, Julie E. Powers, Maya L. Nadimpalli, Timothy R. Julian, Amy J. Pickering

**Author notes:** These authors contributed equally to the work (Abigail P. Harvey, Erica R. Fuhrmeister).

## Abstract

Environmental surveillance of surface contamination is an unexplored tool for understanding transmission of SARS-CoV-2 in community settings. We conducted longitudinal swab sampling of high-touch non-porous surfaces in a Massachusetts town during a COVID-19 outbreak from April to June 2020. Twenty-nine of 348 (8.3 %) surface samples were positive for SARS-CoV-2, including crosswalk buttons, trash can handles, and door handles of essential business entrances (grocery store, liquor store, bank, and gas station). The estimated risk of infection from touching a contaminated surface was low (less than 5 in 10,000), suggesting fomites play a minimal role in SARS-CoV-2 community transmission. The weekly percentage of positive samples (out of *n*=33 unique surfaces per week) best predicted variation in city-level COVID-19 cases using a 7-day lead time. Environmental surveillance of SARS-CoV-2 RNA on high-touch surfaces could be a useful tool to provide early warning of COVID-19 case trends.

## Introduction

Severe acute respiratory syndrome coronavirus 2 (SARS-CoV-2), the virus causing the current global COVID-19 pandemic, is believed to be transmitted primarily through droplets and aerosols.^1^ However, the role of fomites in transmission is unclear.^2^ Recent commentaries argue that the risk of transmission via fomites may be low in clinical settings,^3,4^ although the World Health Organization (WHO) states that fomites may contribute to the spread of COVID-19.^5^ SARS-CoV-2 has been found to remain viable on surfaces for up to 28 days, with half-lives on plastic and stainless steel ranging from hours to days depending on initial concentration and environmental conditions.^6–11^ In clinical settings, detection of SARS-CoV-2 RNA on surfaces is common, particularly in bathrooms.^12–16^ SARS-CoV-2 RNA has also been detected in a limited number of community locations including a nursing home, ferry boat, pharmacy, gas station, city hall, and near hospitals.^17–19^ However, data on the prevalence of SARS-CoV-2 on high-touch surfaces at essential businesses are limited, and temporal trends during a COVID-19 outbreak have not been measured.

Environmental surveillance is an emerging field for monitoring infectious disease prevalence and trends at the population level. Surveillance of environmental reservoirs has the potential to be less invasive, lower cost, and less biased than sampling individuals, particularly for pathogens with a high proportion of asymptomatic infections. Wastewater sampling (also called wastewater-based epidemiology) has successfully been used to track outbreaks that are otherwise difficult to capture through clinical surveillance such as poliovirus and SARS-CoV-2.^20,21^ Recent studies have documented that SARS-CoV-2 RNA levels in wastewater track with trends in case numbers in communities.^22–30^ However, wastewater epidemiology has not yet been demonstrated to be an early warning system for COVID-19 cases.^31^ In New Haven, Connecticut, wastewater surveillance only provided early warning of COVID-19 cases when there was a delay between specimen collection dates and reporting of test results.^24^

Environmental surveillance methods that do not rely on shedding in stool, such as fomite or air sampling, may be better situated to provide early warning of spikes in COVID-19 cases. Viral load in the upper respiratory tract peaks within one week after symptom onset, whereas viral load in stool has been found to peak one to six weeks after symptom onset.^32–40^ Pre- and asymptomatic patients also shed SARS-CoV-2 in the respiratory tract;^41,42^ thus, environmental surveillance may capture trends among total cases.^43^ Targeted sampling of high-touch surfaces has the potential to complement other pandemic surveillance strategies by identifying recent locations (e.g., buildings or rooms) of currently infectious individuals, providing insight on fomite transmission pathways, and serving as an early warning system of case trends.

We collected longitudinal high-touch surface samples in public locations and essential businesses throughout a COVID-19 outbreak from March 13-June 23, 2020 in Somerville, Massachusetts. Our objectives were to: 1) document the types of high-touch nonporous surfaces likely to be contaminated with SARS-CoV-2 during an outbreak, 2) measure the concentration of SARS-CoV-2 on surfaces to estimate risk of infection from contact with fomites in the community setting, and 3) assess the temporal relationship between environmental surface SARS-CoV-2 contamination levels and COVID-19 cases in the community.

## Results

### Detection of SARS-CoV-2 on surfaces

We collected surface swab samples and recorded touches on 33 unique surfaces at 12 locations in Somerville, MA, including a trash can, liquor store, bank, metro entrance, grocery store, gas station, laundromat, restaurant, convenience store, post office box, and crosswalks. We measured SARS-CoV-2 RNA in surface swab samples by real-time quantitative reverse transcription polymerase chain reaction (RT-qPCR) using the N1^44^ and E Sarbeco^45^ assays. Overall, 29 of 348 (8.3 %) total surface samples were positive for SARS-CoV-2 RNA, and we detected SARS-CoV-2 on surfaces in 10 out of 12 locations sampled (Figure 1). Of all surfaces sampled, 17 (52 %) were positive for SARS-CoV-2 at least once. SARS-CoV-2 was detected on surfaces at all locations except for the convenience store and post office box; the percentage of samples positive by location ranged from 0-25 %. A trash can handle and liquor store door handle were the most frequently contaminated surfaces. Among the 29 samples that were positive by the N1 or E assays, only three amplified in all replicates above the limit of quantification (LOQ): the interior and exterior grocery store door handles on June 16 and the liquor store door handle on May 5. Quantities in these samples ranged from 2.5-102 gc/cm^2^ (Table S1).

**Figure 1:**
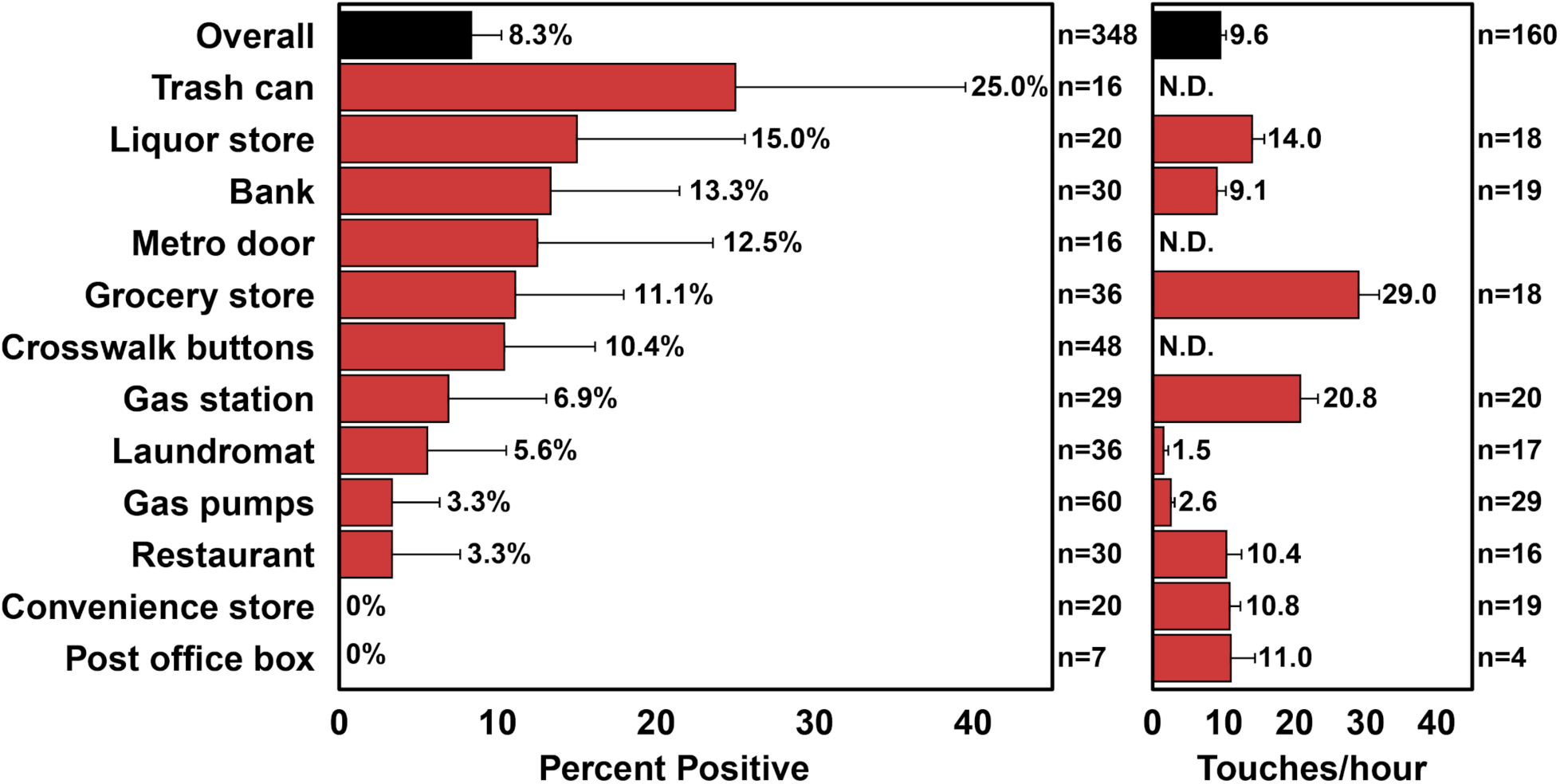
Percentage of positive samples over the duration of the study (left) and mean touches per hour (right) at sampling locations. Error bars show the 90 % confidence interval around the mean. For percent positive, *n*=number of samples collected. For touches/hour, *n*=number of surfaces observed. N.D. signifies that no observational data was collected at that location.

We determined swab recovery efficiencies in the lab by spiking metal and plastic surfaces with a known concentration of bovine coronavirus (BCoV), and also applied BCoV directly to swabs. We recovered 60 % (standard error (SE): 15 %) of BCoV RNA applied directly to swabs, compared to direct extraction. The BCoV RNA swab recoveries were significantly different when swabbing plastic *versus* metal surfaces (*t*=-4.18, *p*=0.02). The overall recovery (compared to direct extraction) was 38 % (4 %) on plastic surfaces and 16 % (2 %) on metal surfaces (Table S3).

The surface area of samples ranged from 1-900 cm^2^ (mean=230 cm^2^, standard deviation (SD)=220 cm^2^). Of the 33 surfaces sampled, 13 were plastic and 20 were metal; there was no significant difference in sample positivity between plastic and metal surfaces (chi-squared test for independence: *χ*^*2*^=0.88, *p*=0.35).

### Longitudinal surface positivity and COVID-19 case trends

We collected samples in two phases: an initial, pilot phase where we sampled 5 unique surfaces twice weekly from March 13-31, 2020, and a full-scale phase, where we sampled 33 unique surfaces at 12 locations weekly from April 23-June 23, 2020. Due to safety restrictions enacted by Tufts University, we paused sampling from April 1-22 which coincided with the peak of new daily cases in Somerville. During the pilot phase from March 13-31, 2020 (*n*=5 surfaces twice weekly), one sample collected on March 27 was positive while all others were negative. The percent of positive samples per week during full-scale sample collection varied from 0-16 %, with peaks occurring on April 28, 2020 and June 16, 2020 (Figure 2). In Somerville, the first COVID-19 case was confirmed on March 4, 2020, and cases peaked on April 10. In the zip code where sampling occurred, peaks in COVID-19 cases occurred on May 5, 2020 and June 16, 2020. We explored lead periods of 0-11 days for the association between sample positivity rate and the 7-day moving average of case numbers, and found that the weekly percentage of positive samples was most strongly associated with COVID-19 cases 7 days later (Figure S1; Table S4). Using the 7-day lead time, the weekly surface positivity rate explained 68.9 % of the variation in COVID-19 cases within the same zip code (*r*=0.83, *r*^*2*^=0.689, *p*=0.003) and 54.8 % of the variation in COVID-19 cases in all of Somerville (*r*=0.74, *r2*=0.548, *p*=0.02; Table S4). Notably, both peaks in surface positivity preceded corresponding peaks in COVID-19 cases within the same zip code by approximately 7 days (Figure 2).

**Figure 2:**
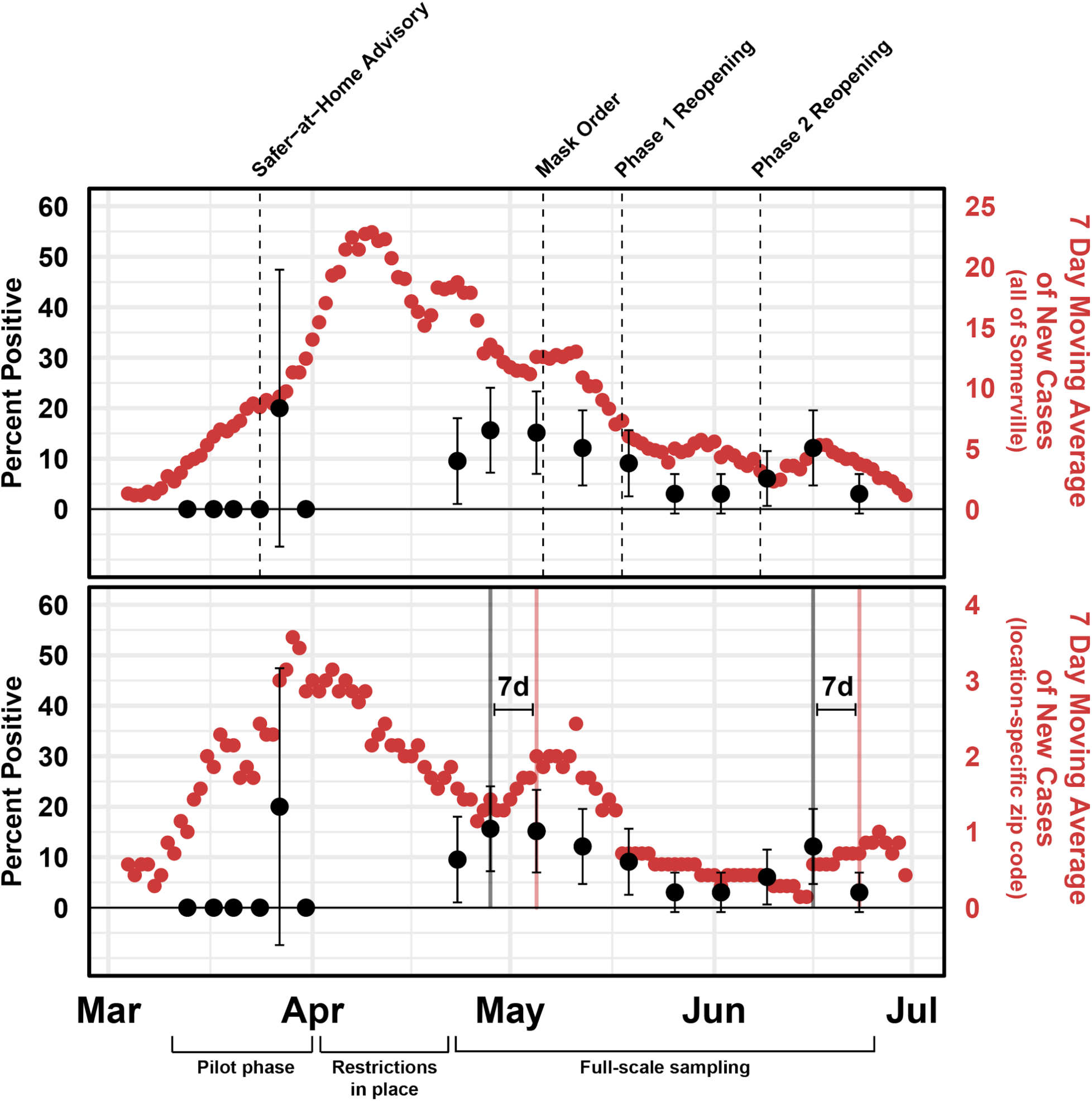
Sample positivity rate and COVID-19 cases. Top: Weekly positivity rate of surface samples and 7-day moving average of new cases in Somerville, MA. The percentage of positive samples are displayed in black and COVID-19 cases in red. Error bars represent the 90 % confidence interval around the percent positive. Sampling was paused from April 1-22 because of restrictions put into place by Tufts University. Bottom: Peaks in percent positivity of surface samples precede 7-day moving average of COVID-19 case peaks in the same zip code by 7 days (shown by vertical black and red lines). On March 24, 2020, a Safer-at-Home Advisory was issued in MA recommending residents shelter in place as much as possible, and all non-essential businesses closed. On May 6, a mask order was issued by the City of Somerville requiring all residents to wear a mask in public spaces. The MA Phase 1 Reopening started on May 18 and allowed some businesses to reopen. The MA Phase 2 Reopening started on June 8 and allowed opening of outdoor dining at restaurants, along with more businesses being allowed to reopen. See mass.gov/info-details/reopening-massachusetts for more details on the MA reopening phases.

### Observational data and sample positivity

We observed a total of 1815 people and 781 bare-hand touches across all sites from April 23 to June 23 (Figure S2). In total, 1623 (89 %) wore masks and 109 (6 %) wore gloves. Of the 977 total touches, 781 (82 %) were bare-hand, 79 (8 %) were gloved-hand, and 117 (12 %) were sleeved-hand. Cloth masks were most common (52 %), followed by surgical masks (40 %), N95 masks (6 %), and other masks (1 %). In Somerville, a mask order was announced April 27, 2020, to be enacted on May 6, 2020; prior to May 5, we observed mask-wearing prevalence at 73 %, and following May 5, mean mask-wearing prevalence increased to 92 % (Figure S2; two-tailed chi-squared independence test: *χ2*=90.6, *p*<0.01). We found no significant time trends in touches or total people present throughout the phases of reopening (Figure S2).

When grouped by location or by week of collection, we found no association between the percentage of positive samples and the number of touches on a surface or number of people per location (Table S5). At the locations with the highest number of touches and visitors per hour (the gas station and grocery store), the percentage of positive samples were not significantly different from the overall positivity rate.

### Temperature and humidity

SARS-CoV-2 survival times on surfaces are known to decrease with rising temperatures.^8,11^ The mean temperature on sampling days was 17 °C (SD 7 °C), the mean relative humidity 61 % (SD 18 %), and mean absolute humidity 13.0 g/m^3^ (SD X; Figure S3). The percent of positive samples per week was inversely associated with daily maximum temperature (Pearson’s *r*=-0.68, *p*=0.03) and absolute humidity (*r*=-0.71, *p*=0.02), while no relationship was found between the percent of positive samples and relative humidity (*r*=-0.44, *p*=0.21). Temperature was also inversely correlated with COVID-19 case numbers (*r*=-0.74, *p*=0.01). Because COVID-19 case trends were correlated with both rising temperatures throughout the spring and a decrease in the weekly percentage of positive samples, we were unable to fully explore the effect of temperature or humidity on SARS-CoV-2 detection rates in this study.

### Infection risk

We estimated the risk of COVID-19 infection from touching a contaminated surface using a quantitative microbial risk assessment (QMRA).^46^ Infection risks ranged from 2 in 10 million to 4 in 10 thousand (mean=6.5×10^−5^, median=2.2×10^−6^). The majority of our positive samples were not quantifiable by qPCR and were therefore treated as the theoretical qPCR limit of detection (LOD) for this analysis. Samples below the limit of quantification had a low risk of infection (2.4×10^−7^ to 3.5×10^−4^), which varied based on object surface area and material (Table S1). Of the three quantifiable samples (door handles at the liquor store on April 28 and May 5 and a grocery store door handle on June 16), the risk ranged from 1 in 100,000 to 4 in 10,000 (Table 1). The QMRA model was most sensitive to the gene copies to infective virus ratio (Spearman’s *r*= −0.53), the dose-response parameter (0.33), the transfer efficiency from surface to hand (0.43), and the transfer efficiency from hand to mucus membranes (0.3).

**Table 1:** Risk of infection from touching sampled surfaces with quantifiable SARS-CoV-2 concentrations.

| Surface | Date | Material | Surface concentration (gc/cm<sup>2</sup>) | Infection Risk |  |  |
| --- | --- | --- | --- | --- | --- | --- |
|  |  |  |  | 5<sup>th</sup> percentile | Median | 95<sup>th</sup> percentile |
| Grocery store door handle | 6/16/20 | metal | 2.54 | $1.82 \times 10^{-6}$ | $1.01 \times 10^{-5}$ | $6.57 \times 10^{-5}$ |
| Grocery store door handle | 6/16/20 | metal | 11.55 | $8.35 \times 10^{-6}$ | $4.66 \times 10^{-5}$ | $3.04 \times 10^{-4}$ |
| Liquor store door handle | 5/5/20 | metal | 102.43 | $7.27 \times 10^{-5}$ | $4.10 \times 10^{-4}$ | $2.60 \times 10^{-3}$ |

## Discussion

We found that the prevalence of SARS-CoV-2 RNA on high-touch surfaces in public spaces and essential businesses reflected, and may even lead, local COVID-19 case numbers by one week. Our findings demonstrate the potential for environmental surveillance of high-touch surfaces to inform disease dynamics during the COVID-19 pandemic. High-touch surface monitoring could be especially useful at finer spatial scales such as within buildings, when regular, widespread human testing is not possible. Surface sampling within buildings could inform the locations of currently infectious individuals and enable early identification of potential COVID-19 cases when individuals are most infectious, including when they are pre-symptomatic or asymptomatic.^41,42^ This may be especially useful within schools or universities, as door entrances and surfaces within each classroom could be tested and, if positive, classrooms could be quarantined before COVID-19 has the chance to spread. An additional advantage of surface surveillance is that swab samples, unlike wastewater samples, can be analyzed using the same sample processing protocols, assays, and biosafety precautions as human nasal swab specimens. As lower cost and rapid diagnostic test assays are being developed during the pandemic for human testing, surface sampling will benefit from these advances as well.^47–51^

Notably, our longitudinal sampling data points are limited (*n*=10 weeks), impacting our ability to definitively prove that surface sampling can be an early warning indicator of case trends. When we included data from the pilot phase (*n*=5 per week), we found no relationship between positive samples and the 7-day moving average of COVID-19 cases in the zip code specific to sampling (*r*=0.33, *p*=0.21) or in all of Somerville (*r*=0.43, *p*=0.10). These results suggest that collecting samples from only 5 surfaces per day was likely not a sufficient sample size in enough locations to capture the relationship between SARS-CoV-2 RNA presence and COVID-19 cases in the community. Further, while only 8.3 % of surfaces were positive for SARS-CoV-2, over 50 % of all surfaces tested positive at least once throughout our study. These results imply that, at the community level, monitoring of one or a few surfaces would not be adequate for environmental surveillance of SARS-CoV-2, and that more widespread sampling in multiple locations may be required to capture true trends in COVID-19 cases. Future research with more frequent sampling in additional settings is needed to confirm the temporal relationship between SARS-CoV-2 presence on community surfaces and COVID-19 cases.

Our findings contribute to a growing literature of detectable but low-level SARS-CoV-2 contamination on public surfaces. Previously reported concentrations of SARS-CoV-2 RNA detected on surfaces in Brazil have ranged from <0.1 - 40 gc/cm^2^ (compared to our 2.5-102 gc/cm^2^) with the fraction of quantifiable to positive samples of 12 % (compared to our 10 %).^17^ The low concentrations detected here and elsewhere may be partially attributable to low recovery of virus RNA from surfaces. Here, recovery was estimated at 16 % for metal surfaces and 38 % for plastic surfaces using BCoV. A previous study also using saline solution and flocked swabs found recoveries of bacteriophage MS2 RNA on surfaces were 9 % on plastic and 7 % on stainless steel, a lesser recovery than ours; however, our recoveries are within the range reported across all eluent-implement combinations (5-40 %).^52^ Future improvements to recovering viral RNA from surfaces could increase assay sensitivity.

Estimated risk of infection from exposure to the contaminated surfaces here is lower than estimates for inhalation exposure to SARS-CoV-2, and lower than fomite transmission risk of other respiratory pathogens. The median risk of infection in our study was lower than the median estimate of infection risk of COVID-19 via aerosols in a seafood market in South China (2.23 x 10^−5^) with only one infected person present.^53^ It is important to note that prior work on aerosol infection risk from SARS-CoV-2 exposure is limited, and risk is dependent on many location-specific factors such as ventilation and number of infected individuals. Compared to other viruses, the risk of fomite-mediated infection in this study is lower than risk of fomite-mediated infection of influenza (median risk=1.25×10^−4^), which is thought to spread primarily via droplets and aerosols,^54,55^ and much lower than risk of norovirus infection (mean risk=2.7×10^−3^), where fomites have been found to play a role in spread.^56–58^

Overall, our results are consistent with the current consensus that fomite-mediated transmission of COVID-19 is possible, but likely a secondary pathway. Our QMRA model was highly sensitive to the ratio of RNA to viable virus; assuming a 1:1 ratio would increase the infectivity risk of our highest-risk sample to a one in 100 chance. We did not attempt to culture live virus from any of our surface samples and therefore cannot determine the viability or infectivity of the SARS-CoV-2 detected in our samples. Future work is needed to confirm the relationship between SARS-CoV-2 RNA concentrations and viable virus on surfaces (which substantially influences the estimated probability of infection) and to determine if infective SARS-CoV-2 can be recovered from fomites in community settings.

Uncertainty in key QMRA model parameters could lead to an underestimate or overestimate of risk. Notably, we estimated risks for a single touch on a single surface. While the risk of an individual transmission event is low, we observed a median of 6 touches per surface per hour, which extrapolates to 336 touches per week (assuming similar touch rates 9am-5pm daily, 7 days per week). Therefore, disinfection of frequently touched surfaces, such as door handles to essential businesses, is likely still useful to prevent possible cases of fomite transmission. Hand disinfection after touching public surfaces could further reduce transmission risk.^46^ Nevertheless, the low infection risk estimated in this study supports prioritizing COVID-19 pandemic response resources to focus on reducing spread via aerosols and droplets (e.g., wearing masks) and by close contacts (e.g., social distancing).

## Methods

### Sample collection

We collected samples from high-touch surfaces in Somerville, Massachusetts, a city with a population of 81,500, a population density of nearly 20,000 people per square mile, and covering 3 zip codes. We collected samples from high-touch surfaces within one zip code in Somerville in two phases: an initial, pilot phase where we sampled 5 unique surfaces twice weekly from March 13-31, 2020, and a full-scale phase, where we sampled 33 unique surfaces at 12 locations weekly from April 23-June 23, 2020. At each location, we sampled 1-6 surfaces, including indoor and outdoor surfaces. Sampling was paused from April 1-22 because of restrictions put into place by Tufts University. During the pilot phase, we sampled 3 crosswalk buttons, a garbage can handle, and a door handle into a metro station. During the full-scale phase, we expanded sample collection to include essential businesses open throughout the sampling period (grocery store, liquor store, convenience store, gas station, laundromat, bank, and restaurant), in which we sampled door handles, ATM keypads, and gas pump handles (Table S6).

Surfaces were swabbed once per week at a fixed day and time using primarily flocked polypropylene swabs (Puritan Medical Products, Guilford, ME), with the exception of the first two weeks of pilot sampling in March when polypropylene swabs were unavailable and cotton-tipped swabs were substituted. We saturated the swab in 1 mL of 1X phosphate-buffered saline solution (PBS), and then swabbed the entire surface horizontally or vertically depending on the surface, rotating the swab throughout. The swabs were then returned to 1 mL of 1X PBS, stored on ice during sampling and transport, and stored at −80°C until further processing. During full-scale sampling, we collected 33 samples and one field blank per week. The field blank consisted of opening a new swab and placing it in PBS in the same manner as the samples.

### Observational data

To determine the number of touches per hour on each surface, we observed each sampling location once per week for 30 minutes at the time of sample collection (time of observations ranged from 10am-5pm). During the observation period, we counted the number and type of touches (bare hand, gloved hand, sleeved hand, and other) on the surface. We also counted the total number of people observed at each location. For door handles, we counted the total number of people entering and/or exiting; for crosswalks, we counted the people crossing that crosswalk; for locations such as ATMs and gas pumps, the total number of people using the service were counted. Finally, we recorded the proportion of people wearing personal protective equipment (PPE) at each location, including the number wearing face masks, face mask type (N95, cloth, surgical, and other), and number wearing gloves.

### COVID-19 case data

Local COVID-19 case data were obtained from the City of Somerville by zip code.^59^ Cases were reported as confirmed, which indicated detection by a molecular test, or probable, which indicated one of several options: (1) a positive antigen test, (2) a positive antibody test and either COVID-19 symptoms or known exposure to a confirmed case, or (3) COVID-19 symptoms with a known exposure to a confirmed case and diagnosis confirmed by a medical provider. Cases were reported by date of sample collection. We calculated total daily cases by summing confirmed and probable tests, and smoothed data using a 7-day moving mean centered on the date.

### RNA extraction and RT-qPCR

We used the QIAamp Viral RNA Mini Kit (Qiagen, Hilden, Germany) to extract RNA from all samples and eluted samples in 80 μl of Buffer AVE. In accordance with the manufacturer’s instructions, the volume of Buffer AVL and ethanol were scaled up proportionally to 1 mL of sample. We added 10 μl of 10^−2^ dilution of bovine coronavirus vaccine stock (Calf-Guard, Zoetis, Parsippany-Troy Hills, NJ) to each sample prior to RNA extraction as an internal standard. To quantify SARS-CoV-2 RNA in samples, we used the CDC N1^44^ and E Sarbeco assays.^45^ Each plate consisted of triplicates of standard curve points, 25 samples, and a no template control. We also used the BCoV assay to quantify the bovine coronavirus internal standard (Table S7).^60^ The limit of detection at which 50% of replicates amplified (LOD50) for each assay was determined through parallel dilutions to quantities between 3 and 10 gene copies (gc)/5μl.^61^ The LOD_50_ was determined to be below 3 gc/5μl for the E assay and between 5 and 7 gc/5μl for the N1 assay (Table S9). The limit of quantification (LOQ) was determined based on the lowest point on the standard curve in which all replicates amplified, which was 4.4 gc/5μl for the E assay and between 4.4 and 44 gc/5μl for the N1 assay, although we treated the LOQ for N1 as 44 gc/5μl for these results (Table S8). Standard curves were calculated using a linear mixed effects model on data pooled from all plates to account for batch effects (Table S8).^62^ A sample was considered positive if at least one of the triplicates amplified with a Ct below 40 in either the N1 or E assay. This method is consistent with other studies on low abundance pathogen targets in environmental matrices.^63–65^ Additional information on extraction and RT-qPCR methods are available in the supporting information.

### Swab recovery efficiency experiment

Swab recovery efficiency was determined by seeding surfaces with 5 µl of bovine coronavirus stock (Bovilis, Merck Animal Health, Madison, NJ) and comparing recovery between swabbing seeded surfaces and direct extraction of bovine coronavirus. Swab recovery experiments were performed in triplicate on metal and plastic surfaces using the polypropylene flocked swabs. Three 10 cm x 10 cm areas were marked off on each surface, then cleaned with CaviWipes (Metrex, Orange, CA), rinsed with deionized water, then cleaned with 70% ethanol followed by RNAse Away (Thermo Fisher, Waltham, MA). The center of each surface was seeded with 5 µl of the bovine coronavirus stock and allowed to dry completely. The swabs were saturated in 1 mL of PBS, and then we swabbed the entire 100 cm^2^ area once vertically and once horizontally. To determine the loss of RNA through drying and swabbing, we seeded 5 µl of the bovine coronavirus stock in triplicate directly onto swabs saturated in 1 mL of PBS. The experimental swabs were then extracted as described above. We also directly extracted RNA from the stock solution to determine the stock RNA concentration.

### Statistical analysis

Pearson correlation coefficients, t-tests, and chi-squared independence tests reported here are all two-tailed. To examine various lag periods for case numbers, we calculated the Pearson correlation coefficient between percent of positive samples and the 7-day case average, including various lag periods of case numbers by shifting the 7-day moving mean data forward by 0-11 days. The highest *r* value was considered the optimal lag period between surface SARS-CoV-2 contamination and COVID-19 cases data.

### QMRA Model

Risks from contacts were estimated using a Quantitative Microbial Risk Assessment (QMRA) framework.^46^ Briefly, probability distributions for the model input parameters were obtained from published scientific literature or this paper (Table S10). The risk of infection was estimated based on the concentration of SARS-CoV-2 on surfaces assuming a single hand-to-surface contact followed by a single hand-to-face contact. A genome copy to infective virus ratio informed by data on respiratory enveloped viruses was used to convert genome copies to Plaque Forming Units (PFU).^66^ The number of viruses transferred from the contaminated surface to the hand upon contact was estimated using the transfer efficiency of viruses between surfaces and hands.^67^ Viral dose was calculated using the contact surface area between the hand and the face^68^ and the transfer efficiency of the virus from the hand to the mucous membranes.^69^ An exponential dose-response model developed elsewhere with pooled data from SARS-CoV and murine hepatitis virus (MHV) was used to calculate the probability of infection.^70,71^ When samples were positive in both the N1 and E Sarbeco assays, the higher concentration was used for QMRA. Additional information on the QMRA model can be found in the supporting information.

## Data Availability

The Somerville COVID-19 dashboard can be accessed at somervillema.gov/covid19dashboard. The sample positivity and COVID-19 case data, as well as R code to replicate our analysis, can be accessed at https://github.com/abharv52/COVID19_longitudinalsampling.

https://somervillema.gov/covid19dashboard

## Author Contributions

A.H, M.C, J.M.S., M.L.N., J.E.P. collected surface swab samples. A.H., J.M.S., M.C., and E.R.F. conducted RNA extractions and qPCR. A.H. conducted all non-modeling data analysis. A.K.P and T.R.J. performed the modeling and risk assessment. A.J.P and T.R.J. designed and oversaw the study. A.J.P, T.R.J., M.L.N., and E.R.F. obtained funding. All authors contributed to the writing of the manuscript.

## Acknowledgements

We thank Alexandria Boehm for advice on study design, methods, and for analyzing our standards and BCoV stock concentration with droplet digital PCR. We thank Elana Chan for assistance with surface sampling. We thank the City of Somerville for providing us with COVID-19 case data. This work was funded by NSF CBET Grant No. 2028623. M.L.N was supported by NIH award KL2TR002545. E.R.F. was supported by the NSF Postdoctoral Research Fellowships in Biology Program under Grant No. 1906957. Any opinions, findings, and conclusions or recommendations expressed in this material are those of the author(s) and do not necessarily reflect the views of the National Science Foundation. We acknowledge that this work was conducted on the unceded traditional territories of the Wampanoag and Massachusetts tribal nations.

## Supplementary Information

### Methods

#### Trial RNA Extractions

We performed trial swab extractions to determine the best method for RNA swab extractions using the QIAamp Viral RNA Mini Kit (Qiagen, Hilden, Germany), RNeasy PowerSoil Total RNA Kit (Qiagen), and the RNeasy PowerWater Kit (Qiagen). Variations of the QIAamp Viral RNA Mini Kit were performed with and without PEG and with a larger volume of buffer AVL. The RNeasy PowerSoil Total RNA Kit was tested with and without beads. Each trial was performed in triplicate. To best replicate the sampling method, swabs were saturated in 1mL of 1X PBS and spiked with 10 µL of bovine coronavirus vaccine stock (Calf-Guard), then extracted according to each kit protocol. The bovine coronavirus vaccine stock was prepared by resuspending 1 vial of vaccine in 1 mL of buffer. The QIAamp Viral RNA Mini Kit without PEG was identified to have the best performance and was therefore used for all swab sample extractions (Table S11).

#### RNA Extraction Procedure

For swab sample RNA extractions using the QIAamp Viral RNA Mini Kit, the swab and 1 mL of PBS were moved to a tube containing 4 mL of the buffer AVL and spiked with 10 µL of a 10^−2^ dilution of the bovine coronavirus vaccine stock as an internal control. We vortexed the solution then incubated for ten minutes at room temperature, discarded the swab, and proceeded with extracting RNA from the remaining solution. The volume of ethanol was increased proportionally to the increased volume of lysis buffer (4 mL). Extraction blanks were included with each set of extractions. Samples were eluted in 80 µl of Buffer AVE, and RNA extracts were stored at −80 °C. Samples were thawed for RNA extraction and the eluent was re-frozen at −80 °C until analysis by RT-qPCR.

#### RT qPCR methods

Samples were analyzed using the N1^1^ and E^2^ assays with a reaction volume of 20 µl (8.5 µl of nuclease-free water, 1.5 µl of combined primer/probe mix, 5 µl of Quantabio UltraPlex 1-Step ToughMix (4X), and 5 µl of template). Twist Bioscience’s (San Francisco, CA) synthetic SARS-CoV-2 RNA control 2 was used as a standard for the N1 and E assays. The standard curve was prepared from the synthetic RNA stock in RNase/DNase-free water to 6-log concentrations ranging from 0.44 copies/5 μL to 4.4×10^4^ copies/5 μL. Due to lack of access to a ddPCR instrument at the beginning of the study, standard curve points between 1 and 10^5^ copies/5 μL were determined using the Twist Biosciences reported concentration of 10^6^ copies/μL and recalculated after measuring the RNA control using ddPCR. The N1 assay cycling conditions consisted of 10 min at 50 °C, then 3 min at 95 °C, followed by 45 cycles of 95 °C for 3 sec and 55 °C for 30 sec. The E assay cycling conditions consisted of 10 min at 55 °C, 3 min at 95 °C, followed by 45 cycles of 95 °C for 15 sec and 58 °C for 30 sec. Each plate consisted of triplicates of standard curve points, 25 samples, and a no template control.

The BCoV assay^3^ was used to test for the internal control. Each reaction volume was 20 μL and consisted of 11 μl of nuclease-free water, 1.5 μl of combined primer/probe mix, 5 μl of Fast Virus MasterMix, and 2.5 μl of template. Genomic RNA standards extracted from a bovine coronavirus only vaccine (Bovilis, Merck Animal Health, Madison, NJ) were used for standards for the BCoV assay. The QIAamp viral RNA Mini Kit was used to extract genomic RNA from the stock vaccine solution. Using the molecular weight of the genome, the copies of target per μL of stock was determined. The genomic RNA was then diluted in RNase/DNase-free water to the following concentrations: 2× 10^4^ copies/2.5 μL, ×10^3^ copies/2.5 μL, ×x10^2^ copies/2.5 μL, 20 copies/2.5 μL, and 2 copies/2.5 μL. Standard curve dilutions for 10-10^6^ copies/2.5 μL were calculated based on the concentration of virus measured using a qubit fluorometer (Thermo Fisher, Waltham, MA) and recalculated after measuring using ddPCR. Cycling conditions for the BCoV assay were 50 °C for 5 min, 95 °C for 20 sec, followed by 45 cycles of 95 °C for 3 sec and 60 °C for 30 sec. Plates consisted of standards and a no template control run in triplicates and 39 samples run in duplicate.

#### RT-qPCR Inhibition Testing

To test for inhibition, additional samples were collected from high-touch surfaces and RNA was extracted using the QIAamp Viral RNA Mini Kit. RNA extract was spiked with the twist RNA control to a concentration of 10^3^ gc/µL. Each sample of extracted RNA was run in triplicate for each assay at the following dilutions: undiluted, 1:2, 1:5, and 1:10. A sample was considered inhibited if the Cq value was less than the theoretical value by at least one.^4^ We detected inhibition in both the N1 and E assays when using Taqman Fast Virus 1-Step MasterMix (Applied Biosystems, Foster City, CA) and in the N1 assay when using TaqPath 1-Step RT-qPCR Mastermix (the E assay was not tested with this mastermix), but no inhibition was found in either assay when Ultraplex 1-Step ToughMix MasterMix (Quantabio, Beverly, MA) was substituted.

#### Temperature and humidity data

The daily temperature and humidity data were obtained from the NOAA tower at Boston Logan Airport.^5^ We used the maximum daily temperature reported in degrees Fahrenheit converted to Celsius, and the average daily relative humidity. Some studies have suggested absolute humidity may modulate virus survival,^6^ so we calculated absolute humidity on sampling days from the temperature and relative humidity data.

#### Observations

The City of Somerville posted signs on crosswalk buttons by May 11, 2020, stating that crosswalk buttons are automatic and people should not push the buttons. Therefore, we did not observe crosswalk buttons or count touches on these surfaces.

#### QMRA Model

Risks from single hand-to-surface followed by hand-to-face contacts were estimated using the Quantitative Microbial Risk Assessment framework.^7^ The Monte Carlo Method (*n*=50,000 simulations) was used to incorporate the variability and uncertainty of the input parameters. Probability distributions for each parameter were obtained from published scientific literature or from this paper (Table S10).

The risk of infection was calculated for each of the 29 surfaces that tested positive for SARS-CoV-2. The concentration of virus on the surfaces (genome copy number (gc)/cm^2^, Table S10) were converted to a concentration of infective virus [PFU/cm^2^] using a genome copy to infectivity ratio, *GC*: *Inf* (Table S10). For the 26 samples that were positive but not quantifiable by qPCR, the reaction quantity was set to 3 gc/5 μL, the theoretical detection limit by qPCR, and quantities per cm^2^ were calculated from each surface area. The number of viruses transferred from the contaminated surface to the hand upon contact 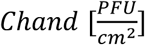 was estimated using the following equation:

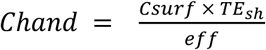

where 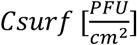 is the concentration of virus in the surface and *TE*_*sh*_[unitless] is the transfer efficiency of viruses between surfaces and hands, and *eff* is the virus recovery efficiency from swabs [unitless]. The dose of virus that enters the susceptible individual through facial membranes, *Dose* [PFU], was estimated as follows:

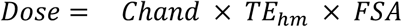

where 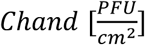 is the concentration of virus in the hand, FSA [cm^2^] is the fractional surface area between the hand and the mucous membranes, and *TE*_*hm*_ [uniteless] is the transfer efficiency of the virus from the hand to the mucous membranes.

The probability of infection, *Pinf*, was estimated using the following exponential model:

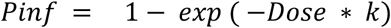

where *k* [PFU^-1^] is the infectivity parameter. The dose-response model can be found in wikiQMRA and is based on the pooled experimental data for SARS-CoV and murine hepatitis virus (MHV)^8,9^.

## Results

All field and extraction blanks (n=36), as well as the no template controls (n=144) included on each qPCR plate, were negative for SARS-CoV-2. Among extraction blanks and field samples, we found similar distributions of RNA extraction yields of the internal standard, bovine coronavirus (BCoV) (Table S2).

The weekly percentage of positive samples explained 56.3 % of the variation in the 7-day moving average of new COVID-19 cases in the zip code specific to the sampling locations (Pearson’s *r*=0.75, *r*^*2*^=.563, *p*=0.01) and 42.3 % in all of Somerville (*r*=0.65, *r*^*2*^=.423, *p*=0.04). The full-scale sampling from April 23-June 23 included sampling on April 23, when only 21 of 33 surfaces from full-scale sampling were collected. Excluding data from this date, the weekly percentage of positive samples explained 60.8% in the zip code specific to the sampling locations (*r*=0.78, *r*^*2*^=.608, *p*=0.01) and 72.3 % of the variation in the 7-day moving average of total COVID-19 new cases in all of Somerville (pearson’s *r*=0.85, *r*^*2*^=.723, *p*=0.04).

A sample was considered positive if it amplified in qPCR in at least one replicate for either the N1 or E assays. If we instead consider samples positive only if they amplify in two or three replicates for either assay, 9 (2.6 %) of samples would be considered positive for SARS-CoV-2.

## Discussion

We found no relationship between the percent of positive samples and the number of touches or number of people when we grouped samples by location or by date. Because SARS-CoV-2 can be eliminated from surfaces through commonly available cleaning solutions,^10^ the cleaning regimen at each sampling location likely impacted SARS-CoV-2 detection. However, we did not monitor the cleaning frequency or method at the sampling locations. For future work, the number of touches since last cleaning may be a more relevant metric to consider for SARS-CoV-2 RNA detection on surfaces.^7^

---

**Figure S1:**
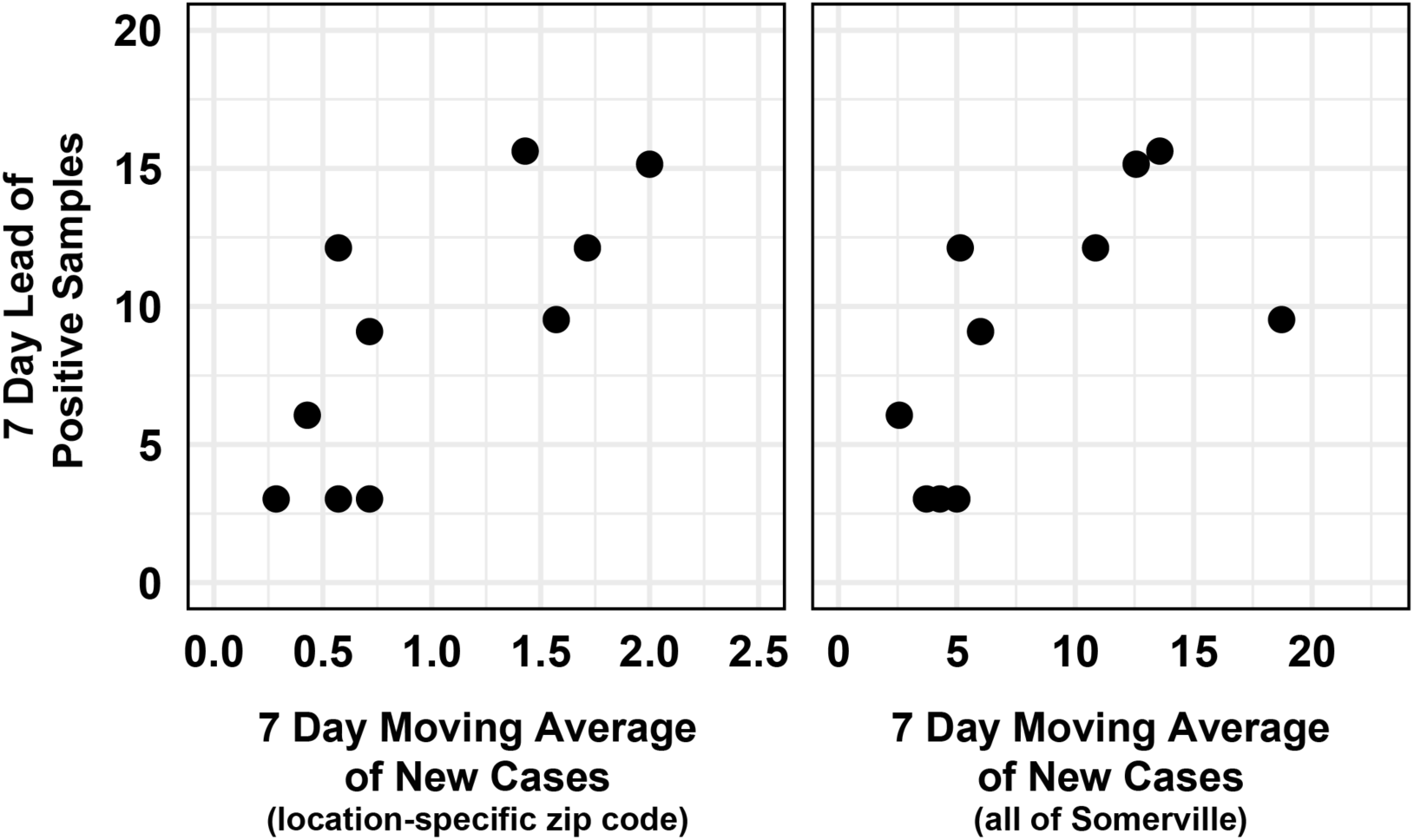
7-day lead of weekly percentage of positive samples (y-axis) versus 7-day moving average of new cases in the location-specific zip code (left) and in all of Somerville (right). The weekly surface positivity rate explained 68.9 % of the variation in COVID-19 cases within the same zip code (*r*=0.83, *r*^*2*^=0.689, *p*=0.003) and 54.8 % of the variation in COVID-19 cases in all of Somerville (*r*=0.74, *r2*=0.548, *p*=0.02).

**Figure S2:**
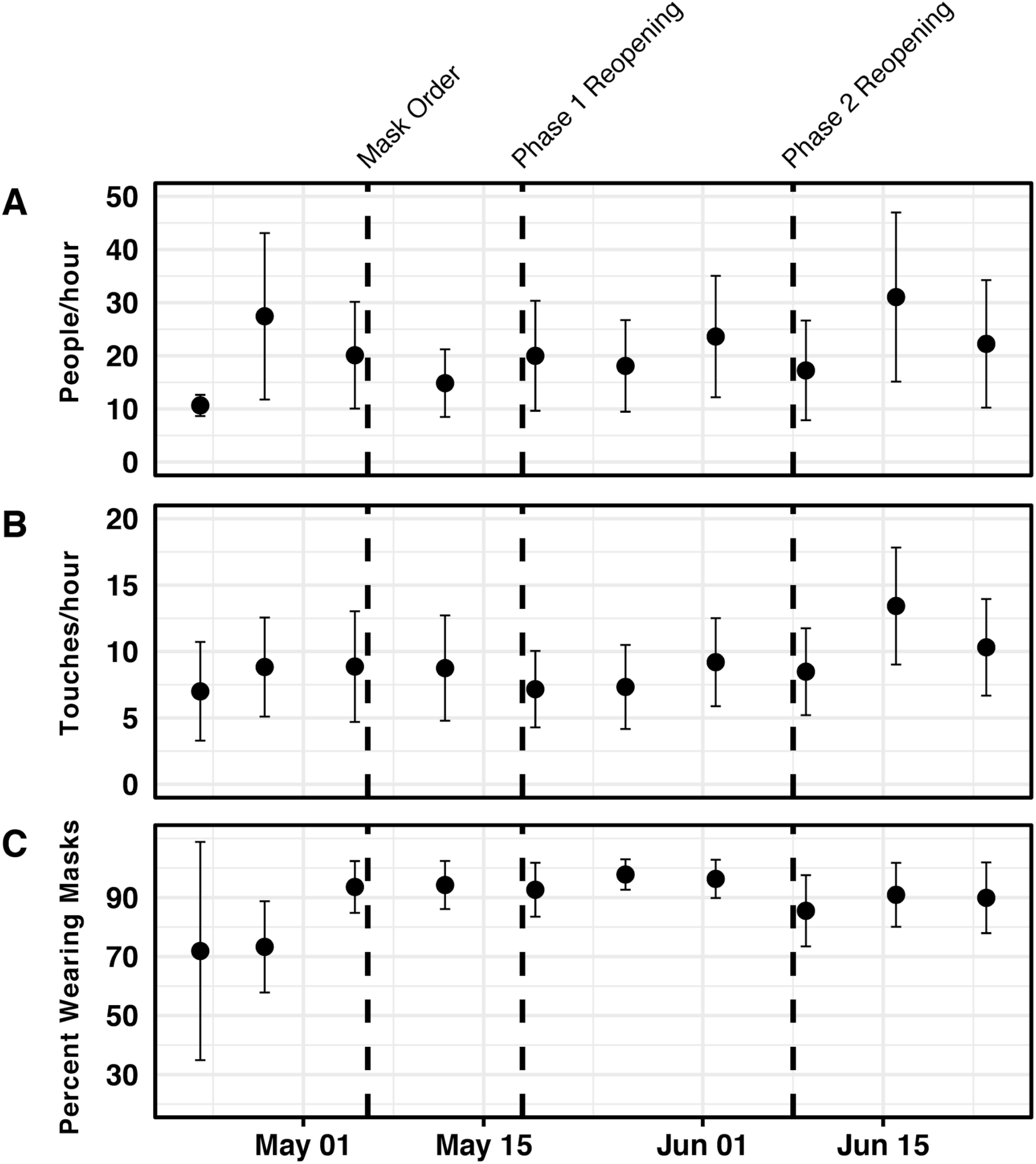
Weekly personal protective equipment prevalence at observation sites April 23-June 23, 2020. A) Mean people visiting observation sites per hour B) Mean touches on sampling surfaces per hour C) Overall percentage of people wearing masks across all sites.

**Figure S3:**
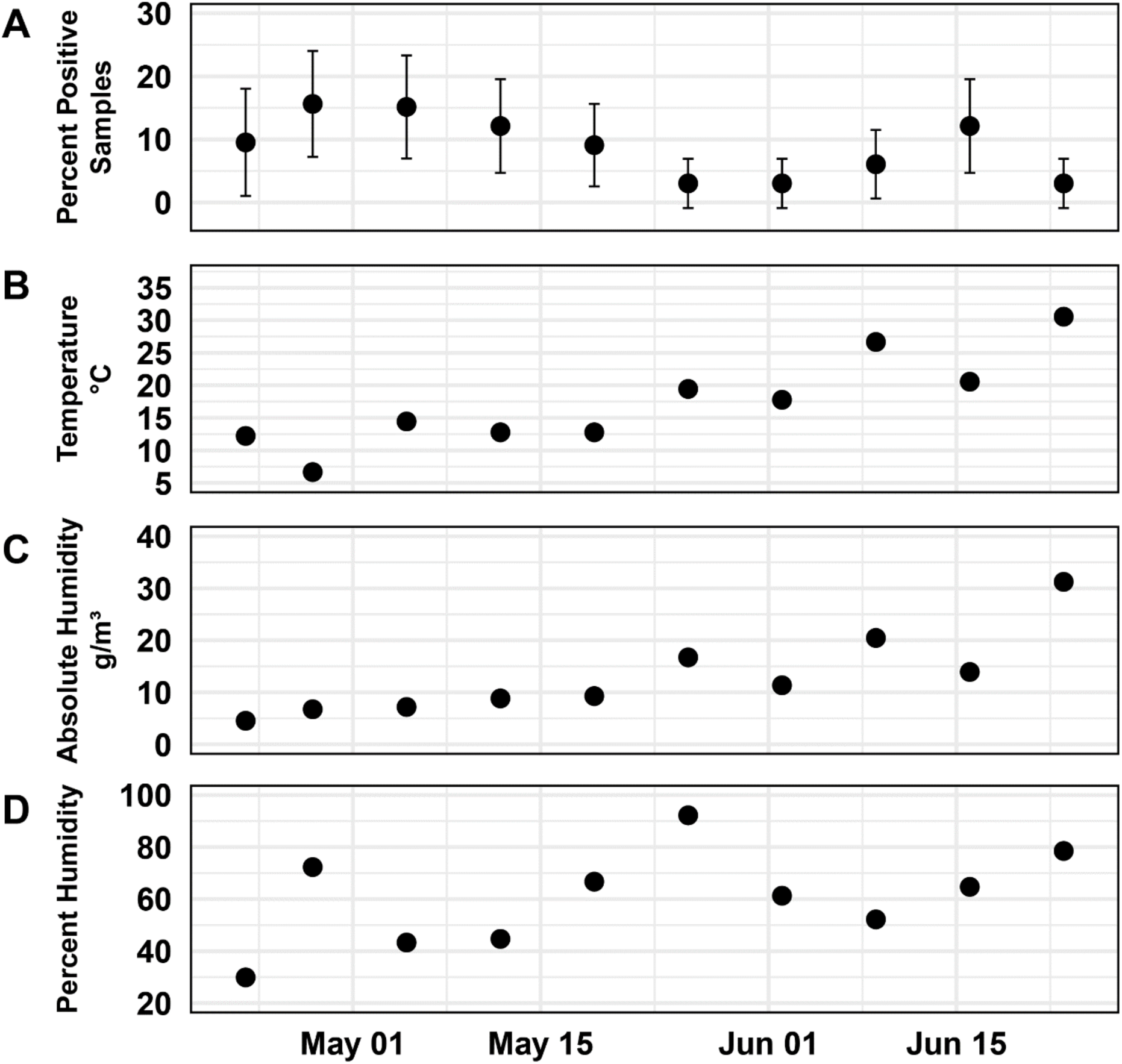
Temperature and relative humidity on sampling days. A) Percent of positive samples per week B) Maximum temperature on sampling days C) Absolute humidity on sampling days D) Average relative humidity on sampling days.

**Table S1:**
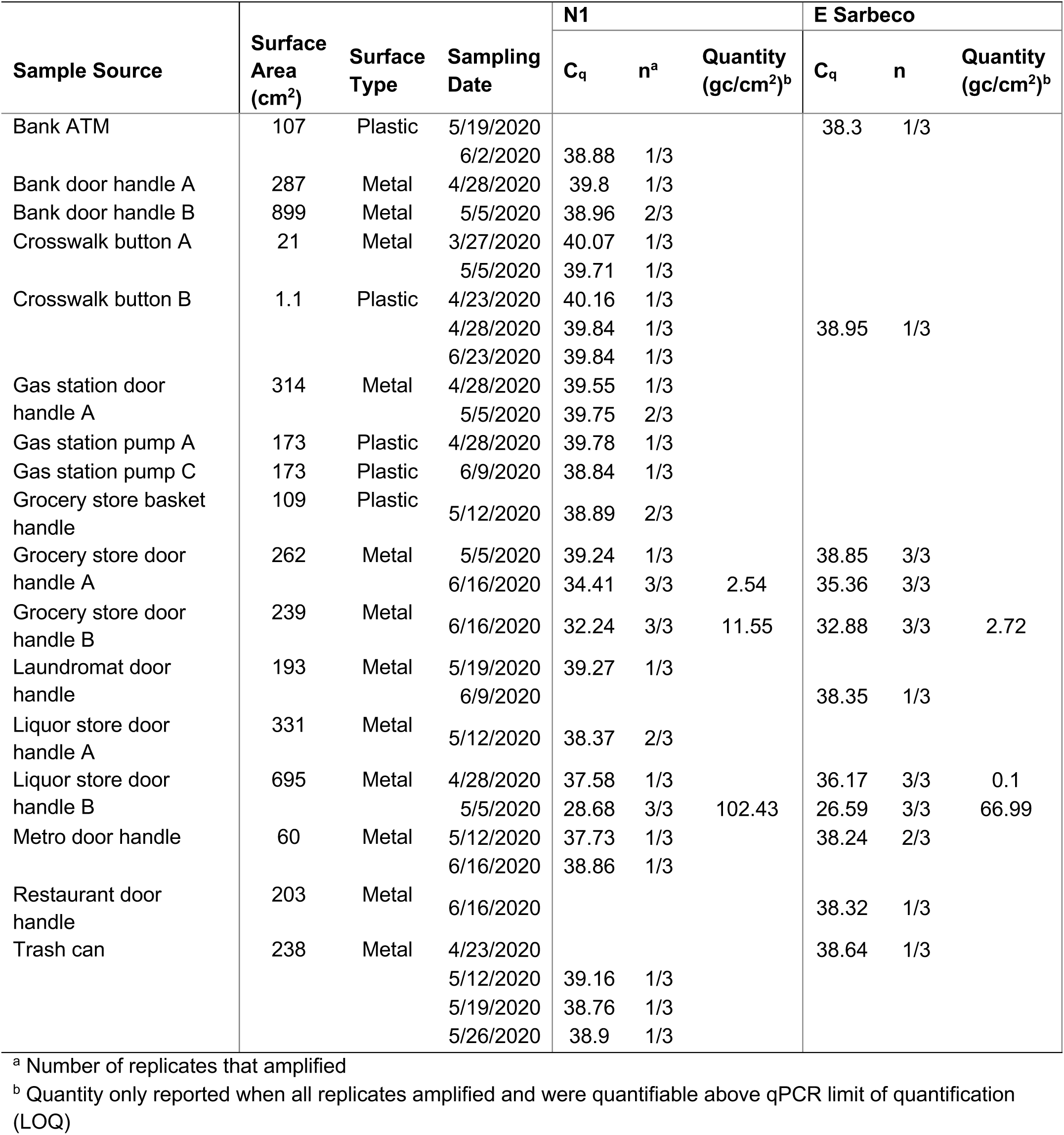
Samples positive for SARS-CoV-2, including surface area of each surface, sampling date, and Ct values and quantities for the N1 and E assays.

| Sample Source | Surface Area (cm<sup>2</sup>) | Surface Type | Sampling Date | N1 |  |  | E Sarbeco |  |  |
| --- | --- | --- | --- | --- | --- | --- | --- | --- | --- |
|  |  |  |  | C<sub>q</sub> | n<sup>a</sup> | Quantity (gc/cm<sup>2</sup>)<sup>b</sup> | C<sub>q</sub> | n | Quantity (gc/cm<sup>2</sup>)<sup>b</sup> |
| Bank ATM | 107 | Plastic | 5/19/2020 |  |  |  | 38.3 | 1/3 |  |
|  |  |  | 6/2/2020 | 38.88 | 1/3 |  |  |  |  |
| Bank door handle A | 287 | Metal | 4/28/2020 | 39.8 | 1/3 |  |  |  |  |
| Bank door handle B | 899 | Metal | 5/5/2020 | 38.96 | 2/3 |  |  |  |  |
| Crosswalk button A | 21 | Metal | 3/27/2020 | 40.07 | 1/3 |  |  |  |  |
|  |  |  | 5/5/2020 | 39.71 | 1/3 |  |  |  |  |
| Crosswalk button B | 1.1 | Plastic | 4/23/2020 | 40.16 | 1/3 |  |  |  |  |
|  |  |  | 4/28/2020 | 39.84 | 1/3 |  | 38.95 | 1/3 |  |
|  |  |  | 6/23/2020 | 39.84 | 1/3 |  |  |  |  |
| Gas station door handle A | 314 | Metal | 4/28/2020 | 39.55 | 1/3 |  |  |  |  |
|  |  |  | 5/5/2020 | 39.75 | 2/3 |  |  |  |  |
| Gas station pump A | 173 | Plastic | 4/28/2020 | 39.78 | 1/3 |  |  |  |  |
| Gas station pump C | 173 | Plastic | 6/9/2020 | 38.84 | 1/3 |  |  |  |  |
| Grocery store basket handle | 109 | Plastic | 5/12/2020 | 38.89 | 2/3 |  |  |  |  |
| Grocery store door handle A | 262 | Metal | 5/5/2020 | 39.24 | 1/3 |  | 38.85 | 3/3 |  |
|  |  |  | 6/16/2020 | 34.41 | 3/3 | 2.54 | 35.36 | 3/3 |  |
| Grocery store door handle B | 239 | Metal | 6/16/2020 | 32.24 | 3/3 | 11.55 | 32.88 | 3/3 | 2.72 |
| Laundromat door handle | 193 | Metal | 5/19/2020 | 39.27 | 1/3 |  |  |  |  |
|  |  |  | 6/9/2020 |  |  |  | 38.35 | 1/3 |  |
| Liquor store door handle A | 331 | Metal | 5/12/2020 | 38.37 | 2/3 |  |  |  |  |
| Liquor store door handle B | 695 | Metal | 4/28/2020 | 37.58 | 1/3 |  | 36.17 | 3/3 | 0.1 |
|  |  |  | 5/5/2020 | 28.68 | 3/3 | 102.43 | 26.59 | 3/3 | 66.99 |
| Metro door handle | 60 | Metal | 5/12/2020 | 37.73 | 1/3 |  | 38.24 | 2/3 |  |
|  |  |  | 6/16/2020 | 38.86 | 1/3 |  |  |  |  |
| Restaurant door handle | 203 | Metal | 6/16/2020 |  |  |  | 38.32 | 1/3 |  |
| Trash can | 238 | Metal | 4/23/2020 |  |  |  | 38.64 | 1/3 |  |
|  |  |  | 5/12/2020 | 39.16 | 1/3 |  |  |  |  |
|  |  |  | 5/19/2020 | 38.76 | 1/3 |  |  |  |  |
|  |  |  | 5/26/2020 | 38.9 | 1/3 |  |  |  |  |

<sup>a</sup> Number of replicates that amplified
<sup>b</sup> Quantity only reported when all replicates amplified and were quantifiable above qPCR limit of quantification (LOQ)

**Table S2:**
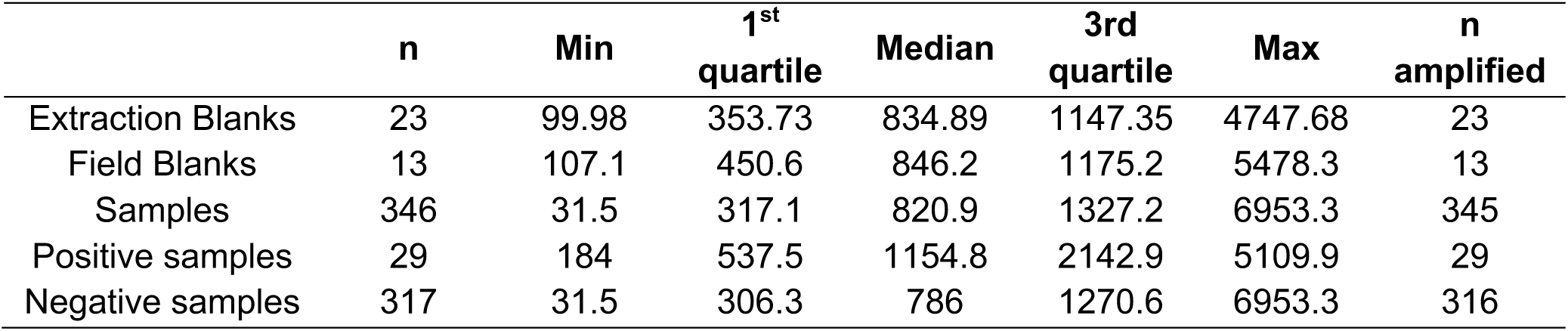
BCoV internal standard recoveries. Quantities are reported in concentration from qPCR reaction (gc/5 µl)

**Table S3:** Results of swab recovery experiments on metal and plastic surfaces compared to direct extraction and direct application to swab.

| **Type** | **Experimental**<br>**Replicates** | **Mean Ct** | **Mean**<br>**Quantity**<br>**(gc/µl)** | **SD**<br>**Quantity** | **Percent**<br>**recovered**<br>**(95% CI)** |
| --- | --- | --- | --- | --- | --- |
| Direct extraction | 2 | 30.22 | 11.54 | 0.07 | - |
| Direct application to<br>swab | 3 | 31.11 | 6.92 | 3.06 | 60.03<br>-15,105 |
| Plastic surface | 3 | 31.69 | 4.39 | 0.74 | 38.03<sup>a</sup><br>(27.3,48.8) |
| Metal surface | 3 | 32.97 | 1.87 | 0.43 | 16.24<br>(9.9, 22.6) |

<sup>a</sup> Percent recovery calculated by comparing recovery on surfaces to recovery by direct extraction.

**Table S4:** Correlation coefficients between COVID-19 cases in the zip code of sample collection and weekly percent of positive samples, lagged between 5 and 11 days.

| **Lag (days)** | **Pearson's r (p-value)** |
| --- | --- |
| 0 | 0.75 (0.01) |
| 5 | 0.80 (0.005) |
| 6 | 0.72 (0.02) |
| 7 | 0.83 (0.003) |
| 8 | 0.82 (0.006) |
| 9 | 0.79 (0.01) |
| 10 | 0.75 (0.02) |
| 11 | 0.70 (0.04) |

**Table S5:** Correlation coefficients between positive samples and observational data: bare hand touches per hour, total people per hour, and percentage of mask-wearers.

| **Category** | **Weekly positive samples (p-value)** | **Positive samples by location (p-value)** |
| --- | --- | --- |
| Bare hand touches/hour | -0.27 (0.46) | 0.33 (0.38) |
| Total people/hour | 0.19 (0.61) | 0.36 (0.34) |
| Percentage of mask-wearers | -0.38 (0.28) | -0.16 (0.64) |

**Table S6:** Locations sampled, type of sample, date of sampling, and number of samples collected.

| Site | Surfaces sampled | Dates of sampling | Total Samples Collected |
| --- | --- | --- | --- |
| Crosswalk buttons | 3 crosswalk buttons | Semiweekly March 13-31 ,2020; Weekly April 23-June 23, 2020 | 48 |
| Trash can | 1 trash can handle | Semiweekly March 13-31,2020; Weekly April 23-June 23, 2020 | 16 |
| Post office box | 1 handle on post office box | Weekly May 12-June 23, 2020 | 7 |
| Metro door | Door handle into metro station | Semiweekly March 13-31,2020; Weekly April 23-June 23, 2020 | 16 |
| Convenience store | 1 exterior door handle, 1 interior door handle | Weekly April 23-June 23, 2020 | 20 |
| Laundromat | 1 exterior door handle, 2 interior door handles, and coin machine dispenser | Weekly April 23-June 23, 2020 | 36 |
| Liquor store | 1 exterior door handle, 1 interior door handle | Weekly April 23-June 23, 2020 | 20 |
| Restaurant | 1 exterior door handle, 1 interior door handle, and door handle to fridge/freezer | Weekly April 23-June 23, 2020 | 30 |
| Bank | 1 exterior door handle, 1 interior door handle, ATM keypads | Weekly April 23-June 23, 2020 | 30 |
| Grocery store | 1 exterior door handle, 1 interior door handle, two grocery carts | Weekly April 23-June 23, 2020 | 36 |
| Gas station | 1 exterior door handle, 1 interior door handle, ATM keypads | Weekly April 23-June 23 | 30 |
| Gas pumps | 3 pump handles and associated keypads | Weekly April 23-June 23 | 60 |

**Table S7:**
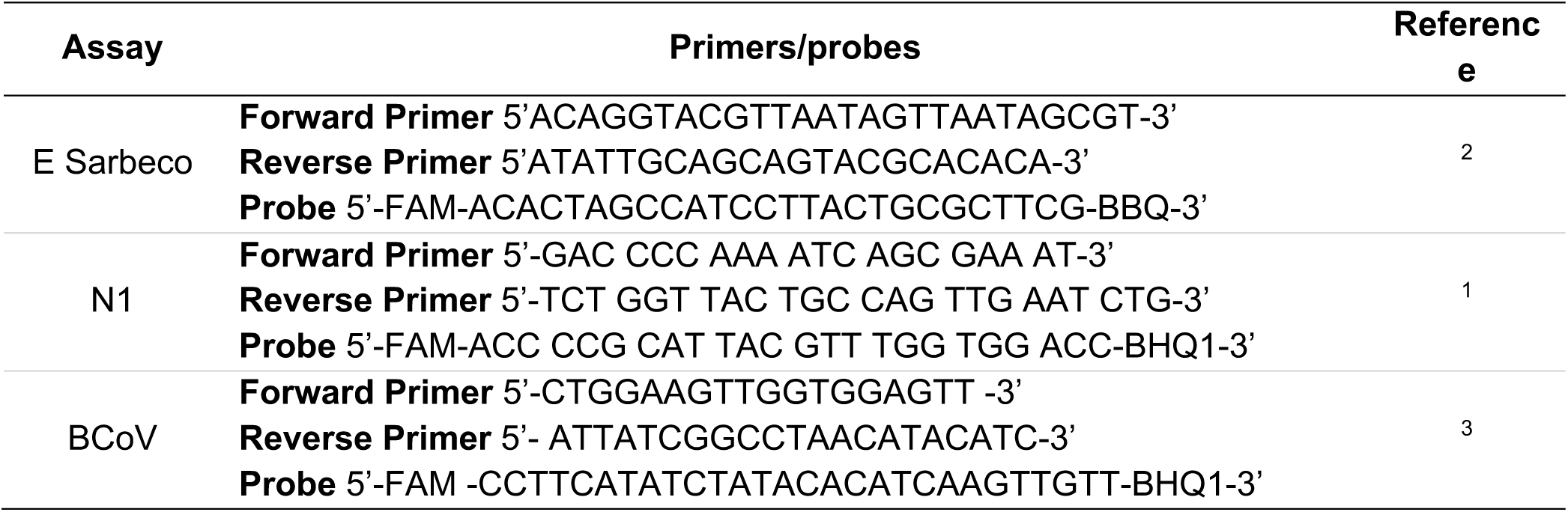
Primers and probes used for RT-qPCR analysis.

**Table S8:** Standard curve slopes, intercepts, efficiencies and R2 for all assays.

| **Assay** | **Slope** | **Mean intercept** | **Efficiency** | **R<sup>2</sup>** | **% Ampl. at 4.4 gc/5ul<sup>a</sup>** | **% Ampl. at 44 gc/5ul** |
| --- | --- | --- | --- | --- | --- | --- |
| N1 | -3.45 | 40.43 | 94.9 | 0.98 | 78 | 100 |
| E | -3.54 | 38.48 | 91.6 | 0.99 | 100 | 100 |
| BCoV | -3.42 | 41.2 | 96.0 | 0.99 |  |  |

<sup>a</sup> % Ampl. signifies the percent of replicates that amplified at that concentration across all standard curves

**Table S9:** Limit of Detection (LOD) results for N1 and E assays.

| **Assay** | **Quantity**<br>**(gc/5µl)** | **n**<br>**amplified** | **Pct<sup>a</sup>** |
| --- | --- | --- | --- |
| **N1** | 3 | 6 | 60 |
|  | 5 | 4 | 40 |
|  | 7 | 7 | 70 |
|  | 10 | 10 | 100 |
| **E** | 3 | 9 | 90 |
|  | 5 | 9 | 90 |
|  | 7 | 9 | 90 |
|  | 10 | 10 | 100 |

<sup>a</sup> Percent amplified out of 10 total replicates

**Table S10:** Probability distributions for model parameters in the Quantitative Microbial Risk Assessment model. Distributions are abbreviated as follows: N = normal (mean, SD); unif = uniform (lower-bound, upper-bound), tri = triangular (min, mode, max).

| Parameter | Description | Distribution | Units | Reference and comments |
| --- | --- | --- | --- | --- |
| GC:Inf | Gene copies to infectivity ratio | unif (100-1000) | Gene copies/<br>PFU | <sup>11</sup> Based on mean gene copy to infectivity ratio for influenza A(N1N1), A(H3N2), and B, the ratio of TCID<sub>50</sub> to PFU, and spare data on SARS-CoV-2<sup>12-14</sup> |
| TE<sub>hm</sub> | Transfer efficiency of virus from hand to mucus membranes | N (0.2, 0.06) | unitless | <sup>15</sup> Transfer of bacteriophage MS2 from hand to saliva |
| TE<sub>sh_stl</sub> | Transfer efficiency of virus between hand and metal | N (0.374, 0.16) | unitless | <sup>16</sup> Transfer of bacteriophage MS2 from steel to hand at a relative humidity of 40-65% |
| TE<sub>sh_pl</sub> | Transfer efficiency of virus between hand and plastic | N (0.795, 0.212) | unitless | <sup>16</sup> Transfer of bacteriophage MS2 from acrylic to hand at a relative humidity of 40-65% |
| FSA | Fractional surface area | unif (3.92, 5.88) | cm<sup>2</sup> | <sup>17</sup> Fractional area of front partial finger<br><sup>18</sup> Average hand surface area for adults |
| eff | Virus recovery efficiency | N (0.60, 0.266) | unitless | This paper. Recovery efficiency from polypropylene swabs |
| k | Dose-response parameter | tri (0.00107, 0.00246, 0.0068) | PFU<sup>-1</sup> | Data obtained from QMRAwiki, based on combined data of <sup>8,9</sup>. The 0.5th, 50th, and 99.5th percentiles were used as min, mode, and max. Intranasal administration of SARS-CoV or MHV-1 to mice. |

**Table S11:**
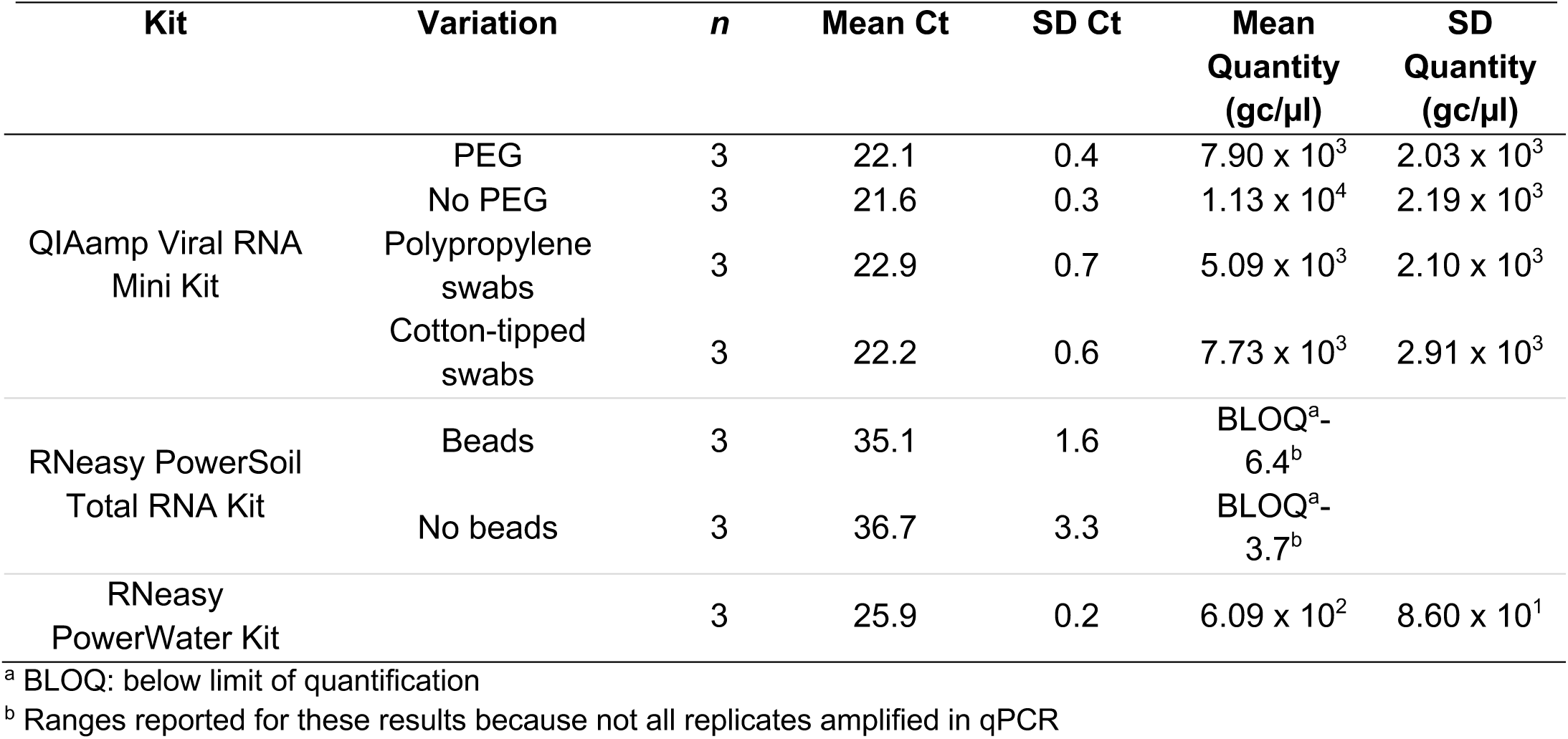
Trial RNA extraction results by extraction kit and protocol variation.

| **Kit** | **Variation** | ***n*** | **Mean Ct** | **SD Ct** | **Mean Quantity (gc/μl)** | **SD Quantity (gc/μl)** |
| --- | --- | --- | --- | --- | --- | --- |
| QIAamp Viral RNA Mini Kit | PEG | 3 | 22.1 | 0.4 | $7.90 \times 10^3$ | $2.03 \times 10^3$ |
| | No PEG | 3 | 21.6 | 0.3 | $1.13 \times 10^4$ | $2.19 \times 10^3$ |
| | Polypropylene swabs | 3 | 22.9 | 0.7 | $5.09 \times 10^3$ | $2.10 \times 10^3$ |
| | Cotton-tipped swabs | 3 | 22.2 | 0.6 | $7.73 \times 10^3$ | $2.91 \times 10^3$ |
| RNeasy PowerSoil Total RNA Kit | Beads | 3 | 35.1 | 1.6 | BLOQ<sup>a</sup>-<br>6.4<sup>b</sup> |  |
|  | No beads | 3 | 36.7 | 3.3 | BLOQ<sup>a</sup>-<br>3.7<sup>b</sup> |  |
| RNeasy PowerWater Kit | | 3 | 25.9 | 0.2 | $6.09 \times 10^2$ | $8.60 \times 10^1$ |

<sup>a</sup> BLOQ: below limit of quantification<sup>b</sup> Ranges reported for these results because not all replicates amplified in qPCR

